# Using the natural language processing system MedNER-J to analyze pharmaceutical care records

**DOI:** 10.1101/2023.09.28.23295887

**Authors:** Yukiko Ohno, Riri Kato, Haruki Ishikawa, Tomohiro Nishiyama, Minae Isawa, Mayumi Mochizuki, Eiji Aramaki, Tohru Aomori

## Abstract

Large language models have propelled recent advances in artificial intelligence technology, facilitating the extraction of medical information from unstructured data such as medical records. Although named entity recognition (NER) is used to extract data from physicians’ records, it has yet to be widely applied to pharmaceutical care records.

In this report, we investigated the feasibility of automatic extraction of patients’ diseases and symptoms from pharmaceutical care records. The verification was performed using MedNER-J, a Japanese disease-extraction system designed for physicians’ records.

MedNER-J was applied to subjective, objective, assessment, and plan data from the care records of 49 patients who received cefazolin sodium injection at Keio University Hospital between April 2018 and March 2019. The performance of MedNER-J was evaluated in terms of precision, recall, and F-measure.

The F-measure of NER for subjective, objective, assessment, and plan data was 0.46, 0.70, 0.76, and 0.35, respectively. In NER and positive–negative classification, the F-measure was 0.28, 0.39, 0.64, and 0.077, respectively. The F-measure of NER for objective and assessment data (F=0.70, 0.76) was higher than that for subjective and plan data, which supported the superiority of NER performance for objective and assessment data. This might be because objective and assessment data contained many technical terms, similar to the training data for MedNER-J. Meanwhile, the F-measure of NER and positive-negative classification was high for assessment data alone (F=0.64), which was attributed to the similarity of its description format and contents to those of the training data.

MedNER-J successfully read pharmaceutical care records and showed the best performance for assessment data. However, challenges remain in analyzing records other than assessment data. Therefore, it will be necessary to reinforce the training data for subjective data in order to apply the system to pharmaceutical care records.

## Introduction

In recent years, with advancements in artificial intelligence technology, it has become possible to extract information related to patients’ diseases and symptoms from unstructured data such as medical records [1, 2].

Technology to extract information such as diseases and symptoms, the names of people and organizations, time expressions, and numerical expressions from text is generally referred to as named entity recognition (NER). Some NER systems also have a positive–negative (P/N) classification function that can be used to determine the onset of extracted findings.

To date, most research on natural language processing (NLP) technology has focused on English texts. NLP technology focused on Japanese texts has lagged due to certain aspects of the Japanese language, including that words are not separated by spaces and subjects are often omitted [3].

Among Japanese NLP studies focused on medical issues, Imai et al. [4] developed a system that performs extraction and P/N classification of malignant findings from radiological reports such as CT reports and MRI reports; Ma et al. [5] built a system that performs extraction and P/N classification of abnormal findings from discharge summaries, progress notes, and nursery notes; and Aramaki et al. [6] developed a system that performs extraction and P/N classification of disease names and symptoms from case history summaries. In addition, Mashima et al. [7] extracted adverse events from progress notes about patients who received intravenous injections of cytotoxic anticancer drugs, and Usui et al. [8] extracted symptomatic states from data stored in the electronic medication records of a community pharmacy and standardized them according to the codes of the International Classification of Diseases, Tenth Revision in order to create a dataset of patients’ complaints. Similar studies have also focused on social media posts and patients’ blogs [9, 10]. The NII Testbeds and Community for Information access Research Project’s “Medical Natural Language Processing for Web Document” task aimed to classify pseudo-tweets according to whether they contained information about patients’ symptoms, and several teams collaborated to build a system to accomplish this task. Nishioka et al. established a system to identify from blog posts whether a patient is positive or negative for hand–foot syndrome on a per-patient and per-sentence basis. Although various approaches have been taken to analyze unstructured medical-related data as described above, most have targeted physicians’ records, including case history summaries, discharge summaries, and radiological reports, NER has not been widely applied to pharmaceutical care records.

Pharmaceutical care records are documents about patients written by pharmacists, who collect information from a pharmacological perspective. Because pharmaceutical care records contain an entry for the change in patients’ physical condition while taking medication, including symptoms of suspected adverse drug effects [11], many such symptoms are documented in pharmaceutical care records. Thus, realizing an NER system that can extract and analyze information from pharmaceutical care records would facilitate investigations of adverse drug effects.

The study by Usui et al. [8], mentioned above, targeted data similar to this study. Because their system was a rule-based model, it had difficulty handling symptoms and contexts that were not set in the rules. Although rules can be added, it is difficult to manage them with consistency. Therefore, we aimed to overcome this problem by using machine learning.

In the present study, we applied MedNER-J, a Japanese-language system designed to extract disease information from physicians’ records [6] to pharmaceutical care records in order to verify the feasibility of NER and P/N classification for this task. Target data were pharmaceutical care records of patients who received cefazolin sodium (CEZ) injection. CEZ is a cephem antibiotics that is often used to prevent secondary infection from operative wounds. The system was applied only to the records of patients who received CEZ injection, with the expectation of mainly collecting target drug information due to fewer concomitant drugs.

## Materials and methods

### Materials

Pharmaceutical care records of patients who received CEZ injection between April 2018 and March 2019 at Keio University Hospital were used as test data (Fig. 1). Researchers accessed and obtained those data on 19 November 2021.

**Fig. 1.**
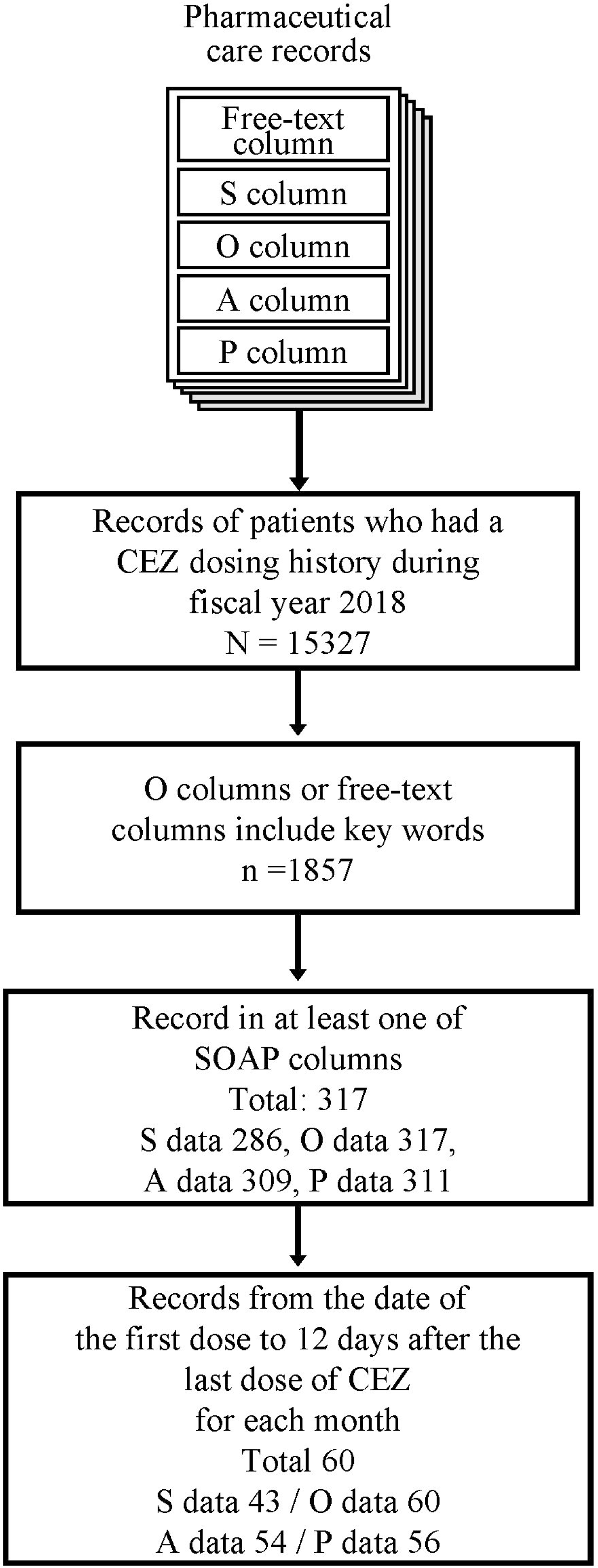
Dataset preparation. Among the records from April 2018 to March 2019, those from the date of first CEZ administration to 12 days after the end of administration that also contained the keywords in the objective column or the free-text column and a record in one of SOAP columns were included in the analysis. S, subjective; O, objective; A, assessment; P, plan.

Pharmaceutical care records were written by pharmacists, and the format consisted of free-text columns and subjective, objective, assessment, and plan (SOAP) columns: subjective information such as patients’ complaints were included in the subjective data; objective information such as clinical history, clinical findings, and laboratory data were included in the objective data; assessments by pharmacists were included in the assessment data; and future plans were included in the plan data.

Data that satisfied the following criteria were used in this research: (1) records with a description in at least one SOAP column, and (2) records including any of the following key words in the free-text column or objective column: cefazolin (written in full-width or half-width katakana characters), cefamezin (written in full-width or half-width katakana characters), CEZ, and cez.

MedNER-J was applied to the records that satisfied the above criteria and that corresponded to the period from first CEZ dosing day to 12 days after the last dosing for each patient for each month.

### Named Entity Recognition / Positive–Negative Classification

We used MedNER-J [12] for NER and P/N classification (Fig. 2). MedNER-J is an NLP system that uses case history summaries as training data and uses conditional random fields [13] based on the feature value of bidirectional encoder representations from transformers [14] to extract diseases and symptoms from physicians’ records. The system can perform P/N classification in order to determine onset or absence of presumed findings from the context.

**Fig. 2.**
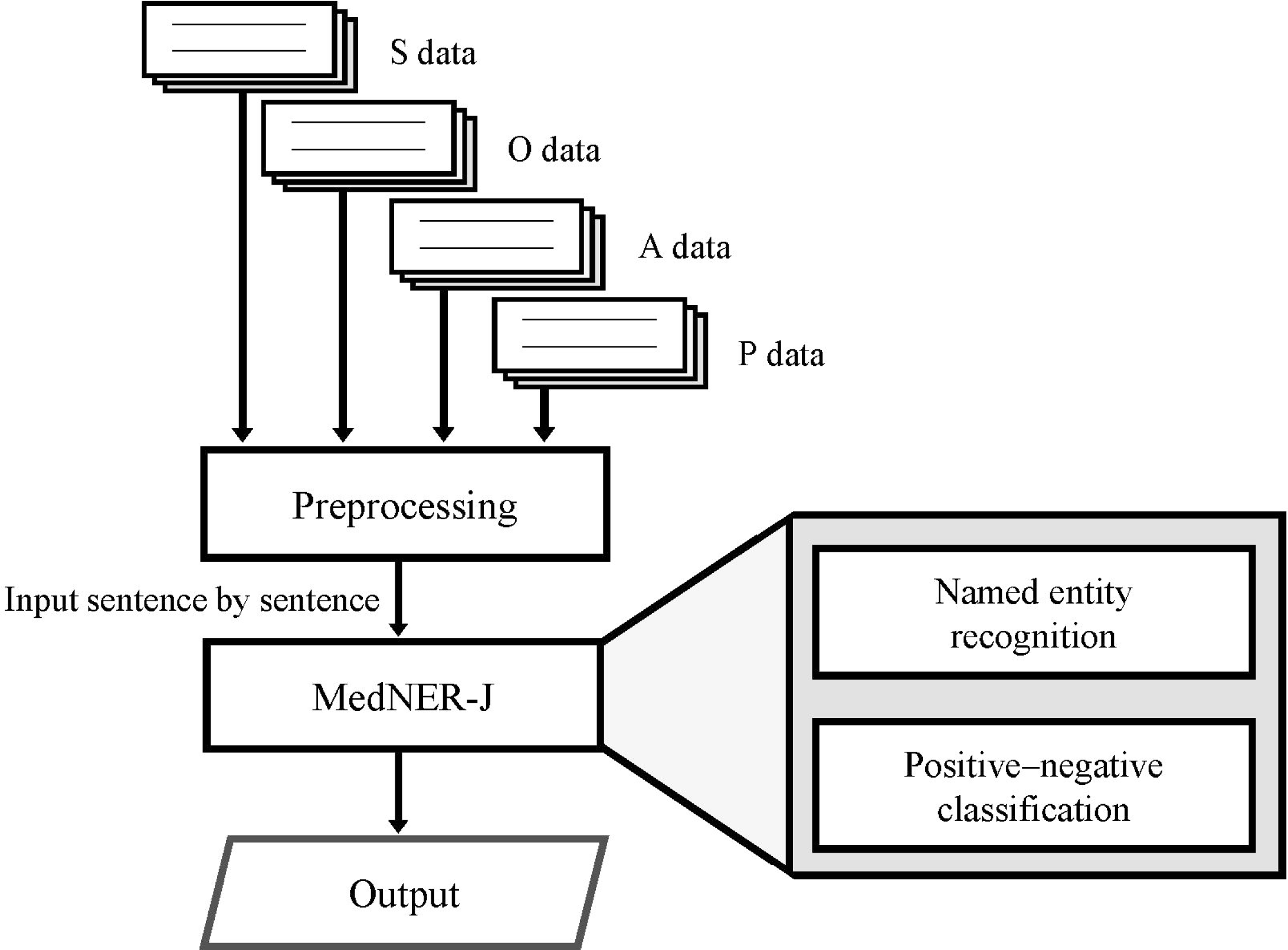
Processing of pharmaceutical care records. Each SOAP column underwent preprocessing as well as NER and P/N classification by MedNER-J in order to obtain the final results. S, subjective; O, objective; A, assessment; P, plan.

As preprocessing, all characters in the records were converted to full-width characters, and exclamation marks were converted to periods.

Preprocessed records were input to MedNER-J on a sentence-by-sentence basis to perform NER and P/N classification. A sentence break was defined as a line break or a period.

### Performance Evaluation

Figure 3 shows the performance evaluation flow. Two researchers independently extracted named entities from the same records, performed P/N classification by visual confirmation, and created the correct answer data. Exact and partial matches of extracted terms between MedNER-J and the two researchers were examined, and P/N classification matches were also investigated. The criteria the researchers followed to create the correct answer data will be explained in the following section.

**Fig. 3.**
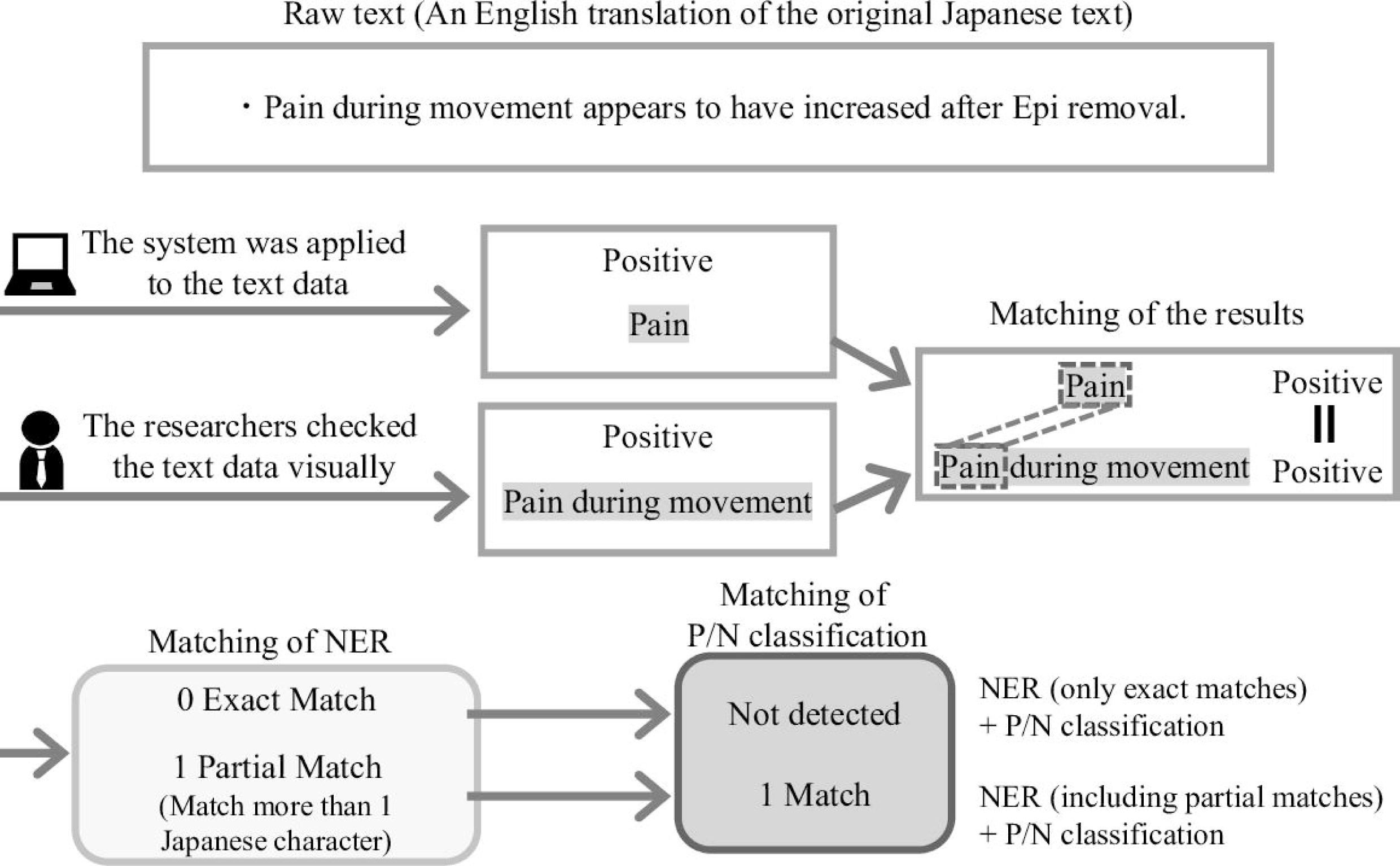
Flow of result matching. The system’s results were matched with the researchers’ results, and performance evaluation indexes were calculated based on the number of NER matches alone and the number of NER and P/N classification matches. Both exact matches as well as partial matches were obtained for NER.

In cases where one sentence contained the same named entities multiple times, researchers also checked whether the positional relationships in the sentence were matched for the same extracted named entities. If the extracted terms matched exactly, they were judged as exact matches. In cases where they did not match exactly but overlapped by one or more Japanese characters, they were judged as partial matches. Both exact match extractions and partial match extractions were checked in terms of P/N classification.

Precision, recall, and F-measure were calculated and evaluated for the following: “matches of NER (including partial matches)” and “matches of NER (including partial matches) and P/N classification.”

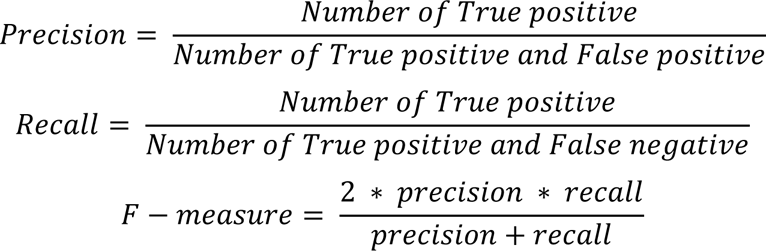

When counting the results, including partial matches, the number of matched terms varied depending on whether they were counted in units of the researchers’ extracted terms or in units of the system’s extracted terms. In such cases, counts were made according to the units, whichever reduced the total number of matched terms.

The validity of researchers’ evaluations was examined using kappa coefficients [15]. Mismatched results between two researchers were discussed and judgement results between researchers were adjusted. The kappa coefficient of the two researchers was 0.87, indicating a high degree of concordance; this showed that researchers’ evaluations were appropriate.

The mismatched results between MedNER-J and the researchers were categorized as follows: (1) system extraction failure, (2) incorrect extraction by the system, (3) difference in P/N classification, and (4) difference in the length of extracted terms. The number of mismatched terms also varied depending on whether they were counted in units of terms extracted by the system or terms extracted by the researchers. In such cases, counts were made according to units, whichever increased the number of mismatched terms.

After categorization, the features of mismatched terms in each category were explored, with the aim of understanding what the system is currently incapable of doing and discussing how those features affect analyses performed by the system.

### Judging Criteria for Researchers

This section outlines the criteria the researchers used to create the correct answer data. Not only nouns such as “pain,” but also verbs such as “hurt,” adjectives such as “sore,” and adverbs such as “painfully” were considered targets for extraction. Symptom modifiers such as site, timing, and severity of symptom onset were also considered together with the symptoms to be extracted. For “sleep,” “appetite,” “state of bowel movements,” “renal function,” “hepatic function,” and “blood electrolyte levels,” if only a statement of normality such as “appetite is fine” was given, it was also considered to be a target for extraction. For example, pharmacists often ask patients whether or not they have experienced a loss of appetite, and patients’ responses, such as “appetite is fine,” were recorded frequently. Such normal states were difficult to consider as diseases or symptoms. Though targets of extraction for records analysis were diseases and symptoms, they are also considered to be important information about patients. Therefore the six items mentioned above were considered for extraction by the researchers. English abbreviations other than laboratory values were consistently excluded from extraction by the researchers. This is because some of them have different meanings among different medical departments, and it was difficult to utilize the extracted terms by themselves. Laboratory values and vital signs were considered for extraction only if words or symbols clearly stated the numerical change or how it was abnormal, with the exceptions of “renal function,” “hepatic function,” and “blood electrolyte concentration.” If only numerical information on laboratory values and vital signs were provided, the information was excluded from extraction because this information is obtainable from the structured data of the medical records, and thus there is no need to extract it from the text data. When symptoms were described consecutively, each symptom was considered as an individual symptom. For “allergy,” any modifiers that indicate the types of allergies listed in the medical dictionary for regulatory activities (MedDRA) was also considered for extraction. For example, if there was a description of “allergy caused by a drug,” this could be classified as “drug hypersensitivity” in MedDRA. Therefore the modifier “caused by a drug” was included in the extracted data. In some cases, specific drug names were mentioned, for example, the description “allergy caused by cefazolin.” However, the drug name “cefazolin” does not appear in MedDRA. If a drug name that does not appear in MedDRA is included in description, only “allergy” was considered as an extraction target, any modifiers were excluded. Although the description “medication for diseases (e.g., diabetes)” was also included, it was not possible to determine whether the medication was used for the patient himself/herself. Therefore, “diseases (diabetes)” in “medication for diseases (diabetes)” was excluded from extraction. “Symptom (e.g., pain)” in “symptom (pain) monitoring” was excluded from extraction because that symptom could not be detected in terms of onset or absence.

In the P/N classification process, the researchers considered symptoms that were currently present in the patients themselves as positive symptoms in principle. Onset or absence of symptoms was determined by referring only to the context within a given sentence. Usage of medication to be taken as needed, such as “times of pain,” was regarded as a negative symptom, because onset has not yet occurred. Adverse drug effects mentioned in the explanation of the drug used were considered to be negative symptoms because they did not actually occur. Past symptoms that were not stated to have resolved, such as “I couldn’t sleep last night,” were considered to be positive symptoms. If there was even a slight improvement in symptoms, they were considered to be negative symptoms. Other cases in which the onset of symptoms could not be determined were considered to be positive symptoms.

### Ethical considerations

This study was approved by the ethics committee of the Keio University School of Medicine (approval No. 2020067). The researchers used only record data that have been previously de-identified by removing patient names and replacing real patient IDs with dummy IDs. Only the personal information manager, who was not included in the authors, had access to the correspondence table between the real patient ID and the dummy ID. The opt-out in written form was implemented instead of informed consent. The opt-out document is available from: http://www.hosp.keio.ac.jp/annai/shinryo/pharmacy/oshirase/.

## Results

Of the 15,327 records of patients who received CEZ injection during the 2018 fiscal year, a total of 317 pharmaceutical care records satisfied both inclusion criteria (Fig. 1). The number of records obtained within the period following CEZ injection were 43 for subjective data (38 patients), 60 for objective data (49 patients), 54 for assessment data (45 patients), and 56 for plan data (46 patients). The number of extracted terms from each SOAP dataset was 50 from subjective data, 411 from objective data, 135 from assessment data, and 37 from plan data by MedNER-J, and 130 from subjective data, 444 from objective data, 216 from assessment data, and 15 from plan data by the two researchers (Table 1). The number of matched extractions, including partial matches between the system and the researchers, was 41 in subjective data, 300 in objective data, 133 in assessment data, and 9 in plan data (Table 1).

**Table 1.**
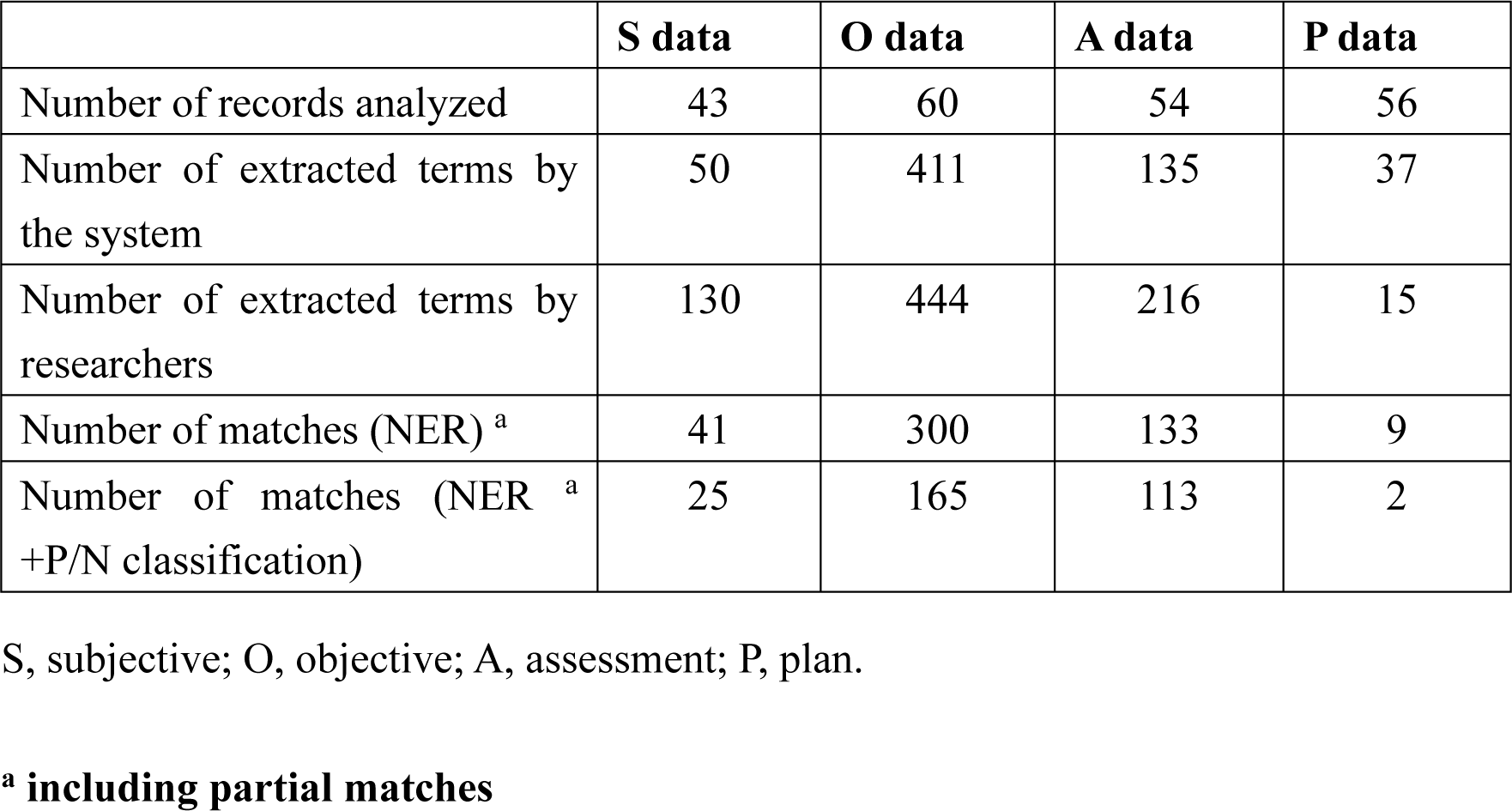
Number of the records analyzed and extraction results by the MedNER-J system and researchers.

The number of terms, for which NER was exactly or partially matched and P/N classification was matched, was 25 for subjective data, 165 for objective data, 113 for assessment data, and 2 for plan data (Table 1).

Table 2 shows the results of the performance evaluation. The precision of NER (including partial matches) was 0.76, 0.82, 0.73, 0.99, and 0.24 for all data, subjective data, objective data, assessment data, and plan data, respectively. The recall of NER (including partial matches) was 0.60, 0.32, 0.68, 0.62, and 0.60 respectively. In NER

**Table 2.** Performance evaluation of NER and P/N classification.

|  | Precision | Recall | F-measure |
| --- | --- | --- | --- |
| NER (including partial matches) |  |  |  |
| All data | 0.76 | 0.60 | 0.67 |
| S data | 0.82 | 0.32 | 0.46 |
| O data | 0.73 | 0.68 | 0.70 |
| A data | 0.99 | 0.62 | 0.76 |
| P data | 0.24 | 0.60 | 0.35 |
| NER (including partial matches) + P/N classification |  |  |  |
| All data | 0.48 | 0.38 | 0.42 |
| S data | 0.50 | 0.19 | 0.28 |
| O data | 0.40 | 0.37 | 0.39 |
| A data | 0.84 | 0.52 | 0.64 |
| P data | 0.054 | 0.13 | 0.077 |

S, subjective; O, objective; A, assessment; P, plan.

(including partial matches) and P/N classification, precision was 0.48, 0.50, 0.40, 0.84, and 0.054, and recall was 0.38, 0.19, 0.37, 0.52, and 0.13 for all data, subjective data, objective data, assessment data, and plan data, respectively. The recall of subjective and assessment data was lower than precision for both NER alone and for NER and P/N classification. Precision was higher than recall for plan data. Recall was similar to precision for objective data.

A trade-off relationship exists between precision and recall, meaning that when one increases, the other decreases. Therefore, the F-measure, which is the harmonic mean of precision and recall, is used as an evaluation index for overall performance. The F-measure of NER alone (including partial matches) was 0.67, 0.46, 0.70, 0.76, and 0.35, while that for NER (including partial matches) and P/N classification was 0.42, 0.28, 0.39, 0.64, and 0.077 for all data, subjective data, objective data, assessment data, and plan data, respectively. These results show that MedNER-J was able to conduct NER and P/N classification with high performance in the order of assessment data, objective data, subjective data, and plan data. Table 3 shows the categories of causes of mismatches between the system and the researchers.

**Table 3.** Percentage of mismatched terms in the total number of extracted terms ^a^ and the number of mismatched terms in each cause category.

| Cause category | S data | O data | A data | P data |
| --- | --- | --- | --- | --- |

|  | [n (%)] | [n (%)] | [n (%)] | [n (%)] |
| --- | --- | --- | --- | --- |
| Total number of extracted terms<sup>b</sup> | 139 | 555 | 218 | 43 |
| **(1) system extraction failure** | 89 (64.0) | 139 (25.0) | 83 (38.1) | 5 (11.6) |
| **(2) incorrect extraction by the system** | 9 (6.47) | 111 (20.0) | 2 (0.900) | 28 (65.1) |
| **(3) difference in P/N classification** | 16 (11.5) | 138 (24.9) | 20 (9.20) | 8 (18.6) |
| **(4) difference in the length of extracted terms** | 13 (9.35) | 81 (14.6) | 30 (13.8) | 2 (4.7) |

S, subjective; O, objective; A, assessment; P, plan.
<sup>b</sup> *Total number of extracted terms =*
*the number of extracted terms by researchers –*
*the number of matched terms between the system and researchers (exact matches and partial match*

Because each type of SOAP data contained differing amounts of information about diseases and symptoms, a comparison of mismatch causes among these data should be based on the percentage of mismatched terms among the total extracted terms (the sum of the number of extracted terms by the system and the researchers minus the number of matched terms), not the number of mismatched terms. In the calculation of this percentage, partial matches were considered matches in cause categories (1) through (3), while partial matches were considered mismatches in cause category (4). For this reason, the percentage of cause categories (1) through (4) does not add up to 100%. Comparing the percentages, the largest percentage of mismatches was subjective data (64.0%) in cause category (1), plan data (65.1%) in cause category (2), objective data (24.9%) in cause category (3), and objective data (14.6%) in cause category (4).

The researchers classified terms in the four cause categories shown in Table 3 into subcategories according to the features of the mismatched term itself and the context around the mismatched term. If a mismatched term had multiple features, it was counted in more than one subcategory.

The subjective and assessment data were expected to contain a large amount of adverse drug effect information due to the characteristics of the SOAP format. The researchers focused on subjective and assessment data because they expected that the analysis of pharmaceutical care records would facilitate the collection and analysis of information on adverse drug effects. Given that the performance for subjective data was low, we listed in Fig. 4 the top five subcategories that had the highest number of eligible cases in cause category (1) with the highest percentage of mismatches in the subjective data.

**Fig. 4.**
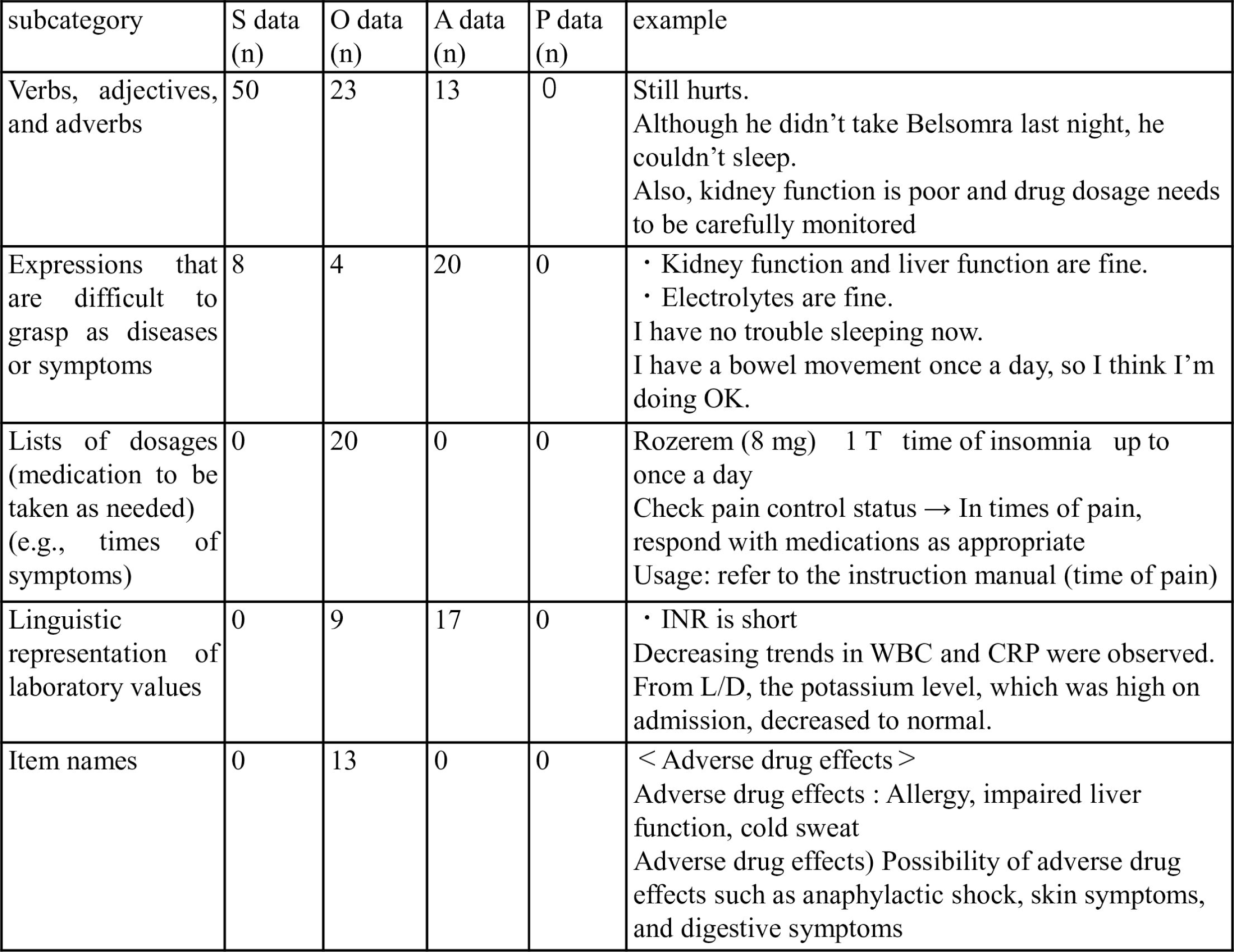
Example breakdown of cause category (1) “system extraction failure.” English translations of the original Japanese texts in the pharmaceutical care records are shown in example. Shading in the example text indicates the scope of the researchers’ extraction. S, subjective; O, objective; A, assessment; P, plan.

The common mismatches in cause category (1) “system extraction failure” were “verbs, adjectives, and adverbs,” “expressions that are difficult to grasp as diseases or symptoms,” and “lists of dosages (medication to be taken as needed) (e.g., times of the symptoms).” The most common mismatches in the subjective data were “verbs, adjectives, and adverbs.”

In mismatches of “verbs, adjectives, and adverbs,” many expressions were general terms or colloquialisms that could be included in the patients’ speech, such as “sore” and “I couldn’t sleep.” The mismatches of “expressions that are difficult to grasp as diseases or symptoms” corresponded to expressions such as “bowel movements are fine.” Although they characterized a normal status, they were important for understanding the patient’s health status. “Lists of dosages (medication to be taken as needed) (e.g., times of the symptoms)” was a description of the dosage of medication to be taken as needed.

## Discussion

### Principal results

Our results showed that when MedNER-J was applied to pharmaceutical care records, NER and P/N classification could successfully be performed. However, the performance of the system differed for each type of SOAP data, and some issues remain for practical utilization. Furthermore, cases in which the system performed inadequately were identified by analysis of mismatch cause categories.

### Application to Pharmaceutical Care Records

The number of extracted terms by both the system and the researchers were larger in the order of objective, assessment, subjective, and plan data. The number of extracted terms by the researchers from each type of SOAP data was 130, 444, 216, and 15 terms, respectively. Furthermore, 41, 300, 133, and 9 terms respectively matched with NER alone (including partial matches) between the system and the researchers. Meanwhile, 25, 165, 113, and 2 terms matched with NER (including partial matches) and P/N classification by the system and the researchers.

The pharmaceutical care records that were targeted in this study included an average of 13.4 diseases or symptoms per record. From these records, MedNER-J correctly extracted an average of 8.1 terms or correctly extracted and performed P/N classification on an average of 5.1 terms. Therefore, MedNER-J was able to extract 60.4% of findings from the pharmaceutical care records and correctly classify 63.0% of those findings as positive or negative.

### Performance evaluation

In this study, we focused on results that included not only exact matches but also partial matches between MedNER-J and the researchers. Word segments in Japanese are unclear, and the necessary extraction range of words varies depending on the situation and the reader. As an example of variations, for the term *itakute* (“in pain”), it is sufficient to extract *itaku* or it may be necessary to extract *itakute*, including the conjunctive particle *te*. In addition, we considered whether expressions related to severity should also be extracted. We speculated that enough information would be extracted from partial matches to ascertain diseases and symptoms. Therefore we decided to analyze results including partial matches.

Although the F-measure for all data was 0.67 for NER alone, and 0.42 for NER and P/N classification, values varied among the subjective, objective, assessment, and plan data. This variation indicates that the applicability of the system differs for each dataset. The F-measure of NER for the objective and assessment data was high (F=0.70, 0.76), while that of NER for the subjective and plan data was only 0.46 and 0.35, respectively. This indicates that the NER performance for the objective and assessment data was superior to that for the subjective and plan data. At the same time, the F-measure of NER and P/N classification was high only for assessment data (F=0.64).

The training data for MedNER-J consisted of case history summaries. Because machine-learning systems are generally optimized for the analysis of the training data, the system was optimized for the analysis of case history summaries. Case history summaries include chief complaints, medical history, laboratory findings, and discussions of each case, as summarized by physicians. Thus, in case history summaries, unlike the pharmaceutical care records written in the SOAP format, the patients’ raw statements in the subjective data could have been replaced by the physicians’ expressions. In addition, the plan data used in this study contained only 15 terms of symptoms, and many records ended with brief descriptions such as “observe the progress.” These points are considered to differ from case history summaries, which describe follow-up plan along with the discussion. This might have resulted in lower performance for the subjective and plan data. In contrast, the objective and assessment data were written in the pharmacists’ expressions and described diseases and symptoms in technical terminology, which likely contributed to the high NER performance. Moreover, “progress and discussion of the disease” are a requisite part of case history summaries [16], and this point was similar to the description of the assessment data. This is probably why the F-measure including P/N classification for the assessment data was high. A decrease in recall implies an increase in false negatives, while a decrease in precision implies an increase in false positives. Therefore, the lower recall than precision for the subjective and assessment data indicate that many mismatches were due to cause category (1) “system extraction failure” in Table 3. In contrast, the lower recall than precision in the plan data indicate that cause category (2), which are incorrect extractions by the system, was more common. In the objective data, recall showed similar values to precision, which means that false positives and false negatives occurred equally without bias.

### Mismatch Cause Subcategories

This section discusses possible failures when the system is used in practice for analysis of pharmaceutical care records, based on the features frequently observed in the cause subcategories. The discussion here focuses on cause category (1), which was the most common cause of mismatches for subjective data. Fig. 4 shows typical examples of cause category (1), which was further divided into 17 subcategories, including “verbs, adjectives, and adverbs,” “expressions that are difficult to grasp as diseases or symptoms,” “lists of dosages (medication to be taken as needed),” “linguistic representation of laboratory values,” and “item names.”

In cause category (1) “system extraction failure,” many extracted terms are categorized as “verbs, adjectives, adverbs” or “expressions that are difficult to grasp as diseases or symptoms.” In “verbs, adjectives, adverbs,” the system was not supposed to extract general terms, such as “sore,” used by patients. The pharmacist receives the patients’ complaints and clinical information and then describes the patient’s condition and other information in objective and assessment columns, replacing them with technical terminology. However, the system’s inability to extract “verbs, adjectives, and adverbs” might cause the pharmacists to overlook symptoms that they did not consider important. Examples of mismatches for extracted terms in the subcategory “expressions that are difficult to grasp as diseases or symptoms” are terms that are related to the disease state but do not directly indicate the disease state, including normal appetite, sleep, bowel movements, renal function, hepatic function, and blood electrolyte levels (Fig. 4). Such normal findings might be missed due to the system’s inability to extract them. One limitation of investigations involving medical records is inability to determine the actual occurrence of symptoms that are not explicitly documented in the medical records. The extraction of normal findings is also important because information that “status of symptoms was documented but they did not occur” is expected to increase the reliability of the results of medical record investigation.

### Future tasks

Not only for cause category (1) but for the other cause categories as well, the cause of the mismatches between the system and the researchers can be explained by one of the following two factors: the training data for the system did not contain similar expressions, or there was a difference between the criteria the system had learned and the criteria the researchers used in this study. Using the analysis target for which performance is expected to be improved as training data should improve the performance of the system. From a medical safety standpoint, overlooking patients’ information is highly detrimental. Therefore, a high recall is preferable, even if precision decreases somewhat. However, recall was significantly lower than precision for the subjective data (precision=0.82, recall=0.32). Therefore, it is critical to improve recall for the subjective data going forward.

Although the SOAP format used in pharmaceutical care records has been the focus of this study, records are sometimes written in SOAP format by other medical staff, including physicians. Among those records, we referred to the subjective data in pharmaceutical care records because of the differences in the kind of attention paid to patients’ changes in clinical state depending on the profession. For example, physicians follow up with patients extensively from disease diagnosis to treatment. Nurses provide not only treatment but also daily care for patients during their hospitalization. In contrast, pharmacists conduct follow-up with patients from a pharmacological perspective, which inevitably includes asking about the beneficial and adverse effects of medications. Therefore, it can be inferred that the descriptions contained in the subjective data of pharmaceutical care records differs from those contained in the subjective data of records by other medical staff, despite the fact they are both subjective data. Consequently, to implement a system that can also analyze pharmaceutical care records, it is imperative to study the subjective data of pharmaceutical care records rather than those of other medical staff.

### Limitation

A limitation of this study is the small sample size, consisting only of patients who received CEZ injection at a single institution. When the system is applied to data from different facilities or data of patients who used different drugs different results might be obtained due to differences in recording formats, adverse drug effect profiles, characterizations of the patients’ chief complaints, and the perspectives of the health care providers.

### Future Utilization

The possibilities for the use of NER in healthcare are broad and varied, as shown by the various efforts undertaken in previous studies [4-10]. Because pharmaceutical care records contain a large amount of information on adverse drug effects, it should be possible to alert healthcare professionals when symptoms of possible adverse drug reactions are extracted with reference to the attached document information. Although medical safety must always be ensured in clinical practice, there is a limit to what can be undertaken due to limited human resources and heavy workloads. However, MedNER-J is expected to help medical staff avoid overlooking patients’ symptoms and thereby improve medical safety. Another possibility is to use the results obtained from analyzing large records to investigate the frequency of adverse drug effects or to discover unknown adverse drug effects based on real-world data. New discoveries might be obtained from analyzing large amounts of data that were previously unavailable.

## Conclusions

MedNER-J, a system designed to extract information from physicians’ records, was applied to extract data from pharmaceutical care records. The system showed high performance for assessment data, was less reliable for other types of SOAP data. Our results suggest that to more effectively apply the system to pharmaceutical care records, the amount of training data needs to be increased to focus mainly on subjective data, which includes patients’ complaints.

## Data Availability

Data of pharmaceutical care records cannot be shared publicly due to an arrangement with the Ethics Committee. Data requests may be sent to the corresponding author.

## References

1. Aramaki E, Wakamiya S, Yada S, Nakamura Y. Natural Language Processing: from Bedside to Everywhere. Yearb Med Inform. 2022 Aug;31(1):243–253. doi: 10.1055/s-0042-1742510. Epub 2022 Jun 2. PMID: 35654422; PMCID: PMC9719781.

2. Dreisbach C, Koleck TA, Bourne PE, Bakken S. A systematic review of natural language processing and text mining of symptoms from electronic patient-authored text data. Int J Med Inform. 2019 May;125:37–46. doi: 10.1016/j.ijmedinf.2019.02.008. Epub 2019 Feb 20. PMID: 30914179; PMCID: PMC6438188.

3. Katsuki M, Narita N, Matsumori Y, Ishida N, Watanabe O, Cai S, et al. Preliminary development of a deep learning-based automated primary headache diagnosis model using Japanese natural language processing of medical questionnaire. Surg Neurol Int. 2020 Dec 29;11:475. Doi: 10.25259/SNI_827_2020. PMID: 33500813; PMCID: PMC7827501.

4. Imai T, Aramaki E, Kajino M, Miyo K, Onogi Y, Ohe K. Finding malignant findings from radiological reports using medical attributes and syntactic information. Stud Health Technol Inform. 2007;129(Pt 1):540–4. PMID: 17911775.

5. Ma X, Imai T, Shinohara E, Sakurai R, Kozaki K, Ohe K. A Semi-Automatic Framework to Identify Abnormal States in EHR Narratives. Stud Health Technol Inform. 2017;245:910–914. PMID: 29295232.

6. Aramaki E, Yano K, Wakamiya S. MedEx/J: A One-Scan Simple and Fast NLP Tool for Japanese Clinical Texts. Stud Health Technol Inform. 2017;245:285–288. PMID: 29295100.

7. Mashima Y, Tamura T, Kunikata J, Tada S, Yamada A, Tanigawa M, et al. Using Natural Language Processing Techniques to Detect Adverse Events From Progress Notes Due to Chemotherapy. Cancer Inform. 2022 Mar 22;21:11769351221085064. doi: 10.1177/11769351221085064. PMID: 35342285; PMCID: PMC8943584.

8. Usui M, Aramaki E, Iwao T, Wakamiya S, Sakamoto T, Mochizuki M. Extraction and Standardization of Patient Complaints from Electronic Medication Histories for Pharmacovigilance: Natural Language Processing Analysis in Japanese. JMIR Med Inform. 2018 Sep 27;6(3):e11021. doi: 10.2196/11021. PMID: 30262450; PMCID: PMC6231790.

9. Wakamiya S, Morita M, Kano Y, Ohkuma T, Aramaki E. Tweet Classification Toward Twitter-Based Disease Surveillance: New Data, Methods, and Evaluations. J Med Internet Res. 2019 Feb 20;21(2):e12783. doi: 10.2196/12783. PMID: 30785407; PMCID: PMC6401666.

10. Nishioka S, Watanabe T, Asano M, Yamamoto T, Kawakami K, Yada S, et al. Identification of hand-foot syndrome from cancer patients’ blog posts: BERT-based deep-learning approach to detect potential adverse drug reaction symptoms. PLoS One. 2022 May 4;17(5):e0267901. doi: 10.1371/journal.pone.0267901. PMID: 35507636; PMCID: PMC9067685.

11. Ministry of Health, Labour and Welfare; [cited 2022 Dec 27]. Available from: https://www.mhlw.go.jp/content/001075622.pdf.

12. MedNER-J [Internet]. Ujiie S, Yata S; [cited 2022 Dec 27]. Available from: https://github.com/sociocom/MedNER-J.

13. Lafferty J, McCallum A, Pereira F. Conditional random fields: Probabilistic models for segmenting and labeling sequence data. ICML. 2001: 282–289.

14. Devlin J, Chang MW, Lee K, Toutanova K. BERT: Pre-training of Deep Bidirectional Transformers for Language Understanding. NAACL-HLT. 2019: 4171–4186.

15. Cohen, J. (1960). A Coefficient of Agreement for Nominal Scales. Educational and Psychological Measurement. 1960; 20(1): 37–46.

16. The Japanese Society of Internal Medicine; [cited 2023 Jan 23]. Available from: https://www.naika.or.jp/wp-content/uploads/J-OSLER/Tebiki_ByorekiHyoka.pdf.

